# Acetate-producing *Prevotella copri* aggravates schizophrenia behaviors by modulating serotonin metabolism

**DOI:** 10.1101/2025.03.25.25324621

**Authors:** Shuai Ji, Qiuxing He, Mengting Su, Shengwei Wu, Yan Liu, Jie Chen, Lijing Du, Dexian Li, Xinghong Zhou, Yanting You, Anna Zuo, Ying Yang, Baizhao Peng, Jie Zhou, Zihao Jiang, Chuanghai Wu, Xiaohu Chen, Ming Wang, Jieyu Chen, Lin Yu, Tommi Vatanen, Xiaoshan Zhao

## Abstract

**Background:** Schizophrenia is a complex mental disorder with approximately 1% lifetime risk worldwide, while the differential diagnosis based on currently clinical criteria is still difficult. A growing literature has demonstrated the biological interactions between gut microbiome and host brain. The metagenomic features may shed the light on how the gut-brain axis works in schizophrenia.

**Results:** A case-control study including 111 fecal metagenomic samples was performed to selected the schizophrenia-associated microbial species and functional modules in comparison with bipolar disorder. The clinical rating scales of both mental diseases were partially explained by gut microbiome. Among these, the *Prevotella copri* was screened out by an integrated approach. The functional annotation and anaerobic culture further confirmed its acetate-producing ability, which was also supported by the changes in serum acetate of schizophrenia patients. Increasing both engraftment of *Prevotella copri* type strain or acetate intake independently induced schizophrenia-like behaviors in mice. Using transcriptomic and metabolomic analysis, the dysregulation of serotonin metabolism across colon, serum and prefrontal cortex was observed after engrafting *Prevotella copri* type strain. The proteins involved in biosynthesis, transportation and degradation of serotonin were significantly elevated after increasing acetate intake.

**Conclusions:** This study suggests that the acetate-producing *Prevotella copri* selected from clinical metagenome can promote schizophrenia-like behaviors in mice by modulating serotonin metabolism. Larger replication studies and further detailed molecular validation are necessary for the generalizability and future clinical applications of the schizophrenia-specific microbial features.

**Graphical Abstract:** 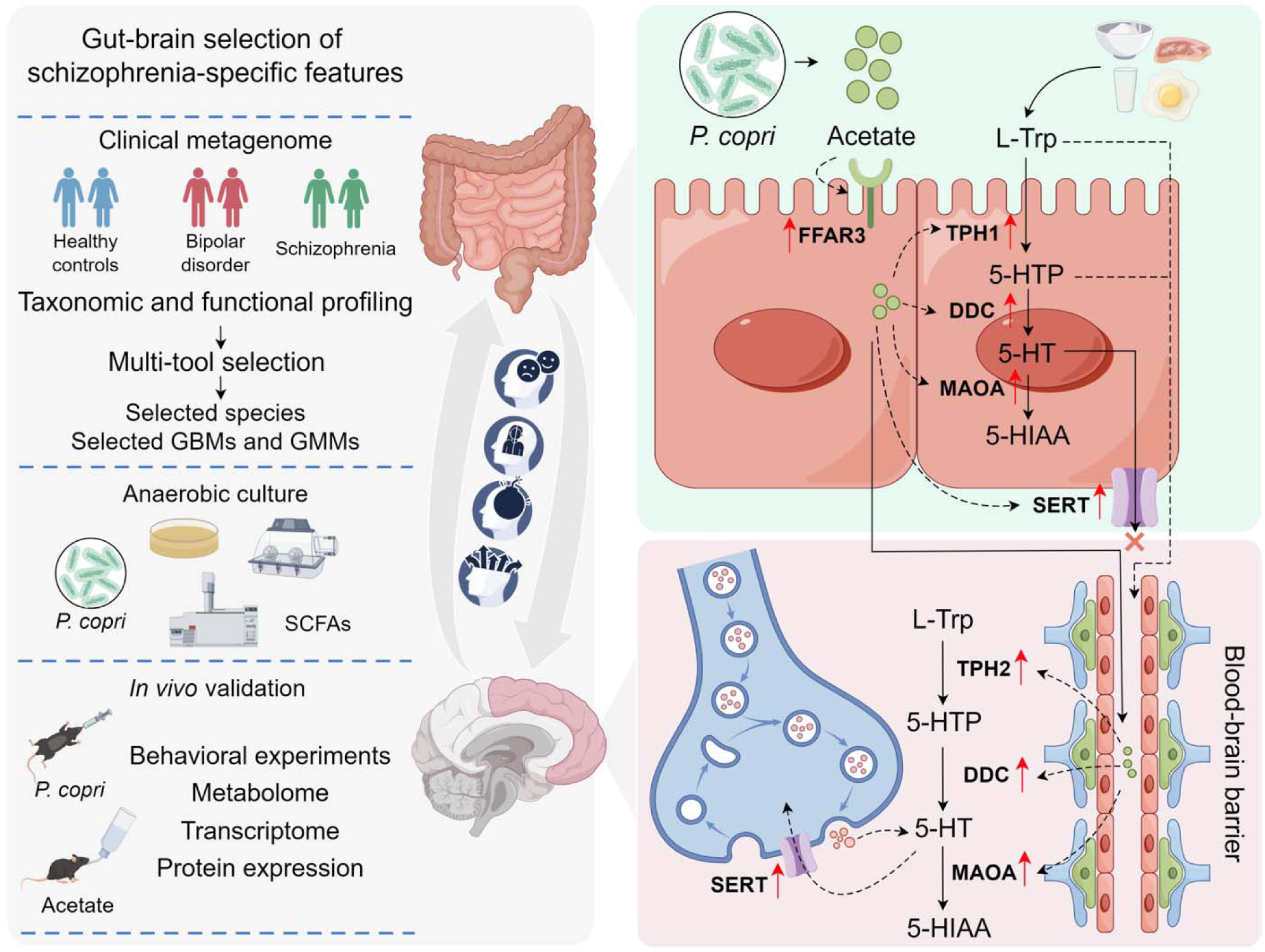

## INTRODUCTION

Mounting evidence suggested the gut microbiome could function as a virtual organ and produced molecules with effects on both the local and distant physiology of the host [1,2]. The biological interactions between gut microbiome and central nervous system were termed the ‘microbiota-gut-brain axis’ [3], suggesting a co-metabolism interaction between gut microbiota and host through substrate co-metabolism and metabolic exchange. Even though the gut microbiome help host maintain health, they can also disrupt homeostasis, influencing the etiology and pathophysiology of mental diseases [4].

As a crucial aromatic amino acid consisting of a β carbon linked to the 3^rd^ position of an indole group, the tryptophan (Trp) is the largest by molecular weight among the 20 common amino acids. Despite its low abundance in protein and cells, the Trp is an important biosynthetic precursor for a wide array of molecules produced by microbiota-host co-metabolism. In the brain, the neurotransmitter serotonin (5-hydroxytryptamine [5-HT]) is produced by the Trp hydroxylase 2 (TpH2). Physiologically, peripheral 5-HT cannot cross the blood-brain barrier, but Trp and 5-HT precursor 5-hydroxytryptophan (5-HTP) can. A majority of 5-HT (over 90%) in the host originates from the gut, especially enterochromaffin cells (ECs). Unlike in the brain, the Trp hydroxylase 1 (TpH1) participates in the biosynthesis of 5-HT in the gut [6]. A previous study has demonstrated 5-HT in the host is mainly regulated by the gut microbiota, indicating low peripheral concentrations of 5-HT and impaired production of 5-HT in the colon of germ-free mice [6]. The short chain fatty acids (SCFAs) have been implicated in stimulating TpH1 expression [7], particularly the acetate [8]. Yet by which specific microbiota and in what changes of this co-metabolism are still unclear.

Globally, the schizophrenia and bipolar disorder (BD) are common severe mental disorders. Around 3% of the general population suffers from BD [9], while 1% suffers from schizophrenia [10]. Most clinical symptoms of patients are interpreted subjectively in the diagnosis, which relies heavily on the training, experience and judgment of the clinician. Worse still, many similarities and commonalities in clinical phenomenology [11], genetics [12] and neuroanatomy [13] have been established between schizophrenia and BD. To date, preclinical and clinical studies have provided evidence supporting the direct or indirect involvement of gut microbiome in schizophrenia and BD [14–16]. Additionally, a psychobiotic diet consisting of more prebiotic and fermented foods has been proposed and proved its efficacy and effectiveness on the reduction of stress and stress-associated mental disorders [17]. A recent case-control study found that functional profiles from metagenomic data, including SCFAs biosynthesis, Trp metabolism and neurotransmitters synthesis/degradation, were associated with the schizophrenia [18]. Two schizophrenia-enriched bacterial species were also selected and transplanted to mice to prove their potential pathogenic roles in schizophrenia, despite missing positive controls [18]. Another clinical study with multi-omics analysis also highlighted the Trp-related neurotransmitter pathways were correlated with the schizophrenia risk, although no in vivo studies were conducted [19]. In this study, we report the schizophrenia-specific metagenomic features compared to BD and healthy controls, including a bacterial species *Prevotella* (*P.*) *copri* and functional modules, through an integrated approach with multivariable methods and a machine learning model. The acetate-related functional module annotated from clinical metagenome were confirmed by anaerobic culture of the type strain of *P. copri* and quantification of SCFAs. The existence of serotonin-related functional modules was further demonstrated experimentally *in vivo* using multi-omics method. The acetate was also shown to independently promote schizophrenia-related behaviors and modulate the biosynthesis, transportation and degradation of serotonin in the colon and prefrontal cortex (PFC).

## RESULTS

### Gut microbial composition and functions distinguish schizophrenia and bipolar disorder

After excluding individuals with baseline variables that may affect intestinal microbiome and host metabolism, serum and fecal samples were collected from healthy controls and patients with schizophrenia and bipolar disorder (Fig. 1a). Detailed information for each individual can be found in Supplemental Table 1. The principal coordinate analysis (PCoA) based on Bray-Curtis distances showed significant differences in gut microbiome taxonomic composition between healthy controls and either disease (Fig. 1b), while did not reveal significant differences between diseases (Supplementary Table 2). Within schizophrenia group, microbial dissimilarity was mainly explained by the depression and excitement score of positive and negative syndrome scale (PANSS) (Fig. 1c). The total score of Hamilton depression rating scale (HAMD) was with the highest contribution to the dissimilarity within bipolar disorder group (Fig. 1d). For host clinical covariates that must be considered, serum iron ions (Fn) and glycosylated serum protein (GSP) may influence gut microbial community in the comparison between two diseases groups (Supplementary Table 2). Nevertheless, the results of microbiome multivariable association with linear models (MaAsLin2) indicated that only two species (*Bacteroides sp. OM05-12* and *Blautia producta*) were affected by the serum levels of Fn and GSP (Supplementary Table 3). To be rigorous, it must also be pointed out that even if schizophrenia patients have not received any antipsychotic treatment for more than three months, the previous therapy time seems to have a lasting impact on the gut microbial communities [20]. Although no specific species were affected by previous therapy time (Supplementary Table 4) and therapeutic modalities (Supplementary Table 5), an additional linear discriminant analysis effect size (LEfSe) algorithm was used to exclude the species with any potential effects of these previous therapeutic modalities (Supplementary Figure 1).

**Fig. 1.**
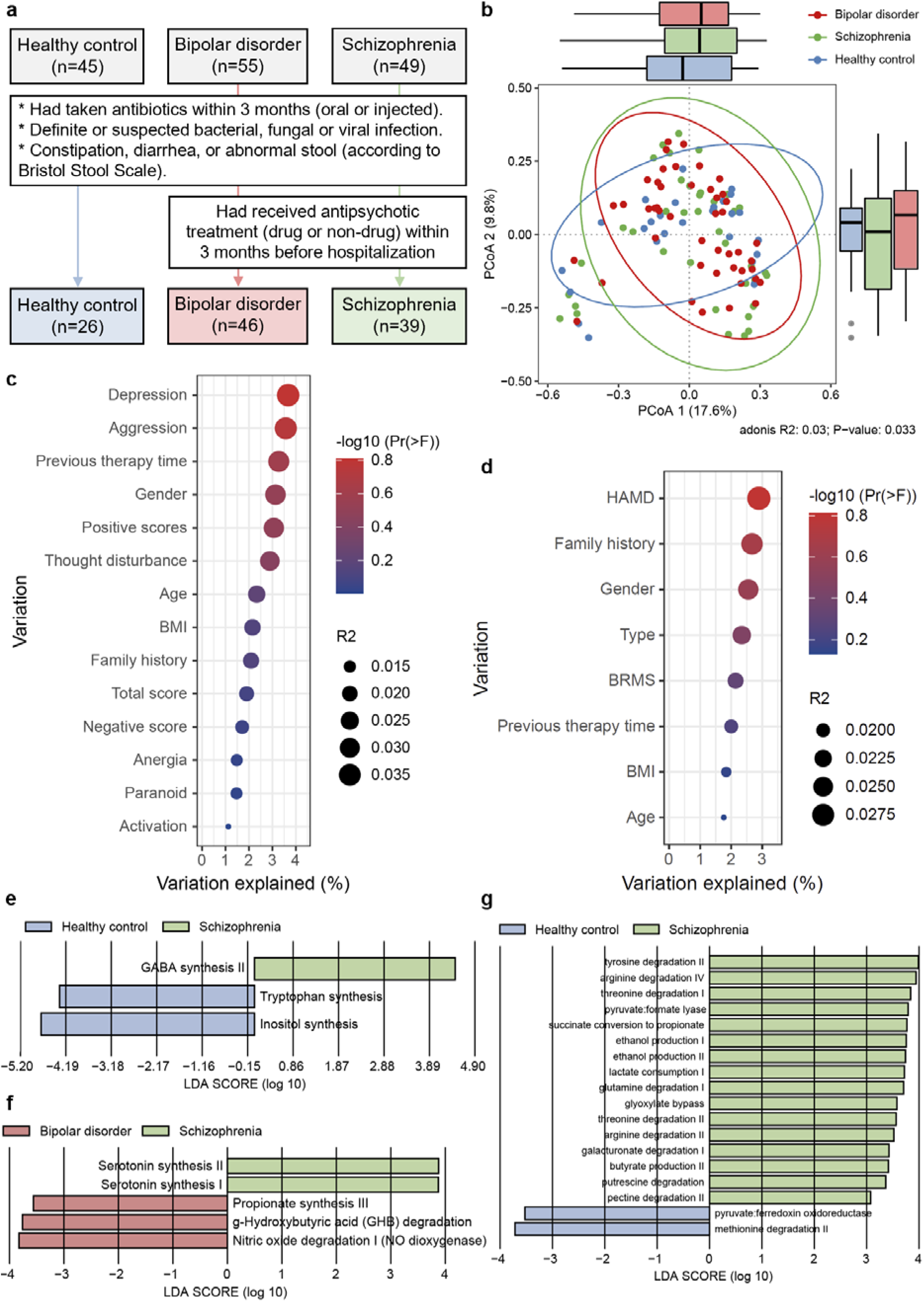
Metagenomic features in patients with schizophrenia or bipolar disorder. **a** An overview of the screening strategy and sample exclusion criteria. **b** The PCoA based on Bray-Curtis dissimilarity between microbiome profiles. Clinical variations of schizophrenia (**c**) and bipolar order (**d**) patients are partially explained by microbiome profiles within each group. **e** LEfSe analysis of GBMs between schizophrenia patients and healthy controls. **f** LEfSe analysis of GBMs between schizophrenia and bipolar disorder patients. **g** LEfSe analysis of GMMs between schizophrenia patients and healthy controls. Significances obtained by LEfSe analysis at p-value < 0.05 and LDA (log_10_) ≧ 2.

For the gut-brain modules (GBMs), Trp synthesis and inositol synthesis in schizophrenia group were significantly lower than that of healthy controls (Fig. 1e). The results were consistent with previous studies [21,22]. Interestingly, the annotated GBMs of serotonin synthesis in schizophrenia group were significantly enhanced compared to bipolar disorder group (Fig. 1f). Among gut metabolic modules (GMMs), the pyruvate formate lyase (PFL) was significantly higher expressed in the gut microbiome from schizophrenia patients (Fig. 1g). The PFL is widely distributed in microbes and contributes to increased ATP production during sugar fermentation in anaerobic environments [23]. With the help of PFL activating enzyme (PFL-AE) and one additional ATP generated from acetyl-CoA conversion, PFL-mediated production of acetate seems to have contributed to higher growth rates and cell yield, which are essential for increasing productivity of the microbial biocatalyst [24]. In acetate environment, PFL can also facilitate the growth of microbes [25]. Other group-wise comparisons of functional modules were depicted in Supplementary Figure 2.

### The acetate-producing species *P. copri* was identified as one of the characteristics of schizophrenia

To select minimal-optimal schizophrenia-specific microbial species, the multivariate methods with unbiased variable selection in R (MUVR) based on a random forest classification model was used and minimal bias variable selection was incorporated into repeated double cross-validation (Fig. 2a). A total of 15 species were selected as strong predictors of the response group, and they were sorted from low to high rank (lower is better) according to the optimal modelling performance (Fig. 2b). These species performed well for distinguishing schizophrenia from bipolar disorder, although the identification of healthy controls appeared to be suboptimal (Supplementary Figure 3). This may be due to the fact that many of these highly abundant species also colonize the intestines of healthy individuals. Linear models in MaAsLin2 were used to further identify which of these species were associated with schizophrenia. With an exclusion of species with any potential effects of clinical laboratory indicators (Supplementary Table 2) and previous antipsychotic therapies (Supplementary Figure 1), *P. copri* was selected as a candidate contributor to schizophrenia (Fig. 2c). Significant linear association was observed between the relative abundance of *P. copri* and the PANSS aggression score within schizophrenia patients (Fig. 1d). As an abundant member of human gut microbiome, *P. copri* may negatively or positively affect host health. There are puzzling discrepancies among studies because *P. copri* strains differ substantially in their encoded metabolic patterns from those of the commonly used reference strain [26]. A previous global meta-analysis separated thousands *P. copri* strains into four distinct clades according to different genetic and population structures [27]. The intriguing dilemma, however, remains that colonizing either too much or too little *P. copri* at the species level seems to be harmful to the gut homoeostasis. Furthermore, gutSMASH algorithm was used to identify primary metabolic gene clusters (MGCs) in the genome of *P. copri* type strain (GCA_000157935.1), revelaving PFL-AE and PFL genes that are involved in the conversion of pyruvate to formate and acetyl-CoA, then generating acetate (Fig. 1e). This MGC exhibited high nucleotide similarity to queried sequences from other species (Fig. 1e) and different strains of *P. copri* (Supplementary Figure 4). The type strain DSM 18205 was cultured *in vitro* to confirm the evidence from functional annotation of clinical metagenome. The concentrations of acetate (Fig. 2f) and butyrate (Fig. 2g) were significantly higher than blank controls after 72 hours of anaerobic incubation. Of note, the serum concentration of acetate in schizophrenia group was significantly higher than both healthy controls and bipolar disorder group (Fig. 2h). There were no significant differences in serum butyrate between groups (Fig. 2i).

**Fig. 2.**
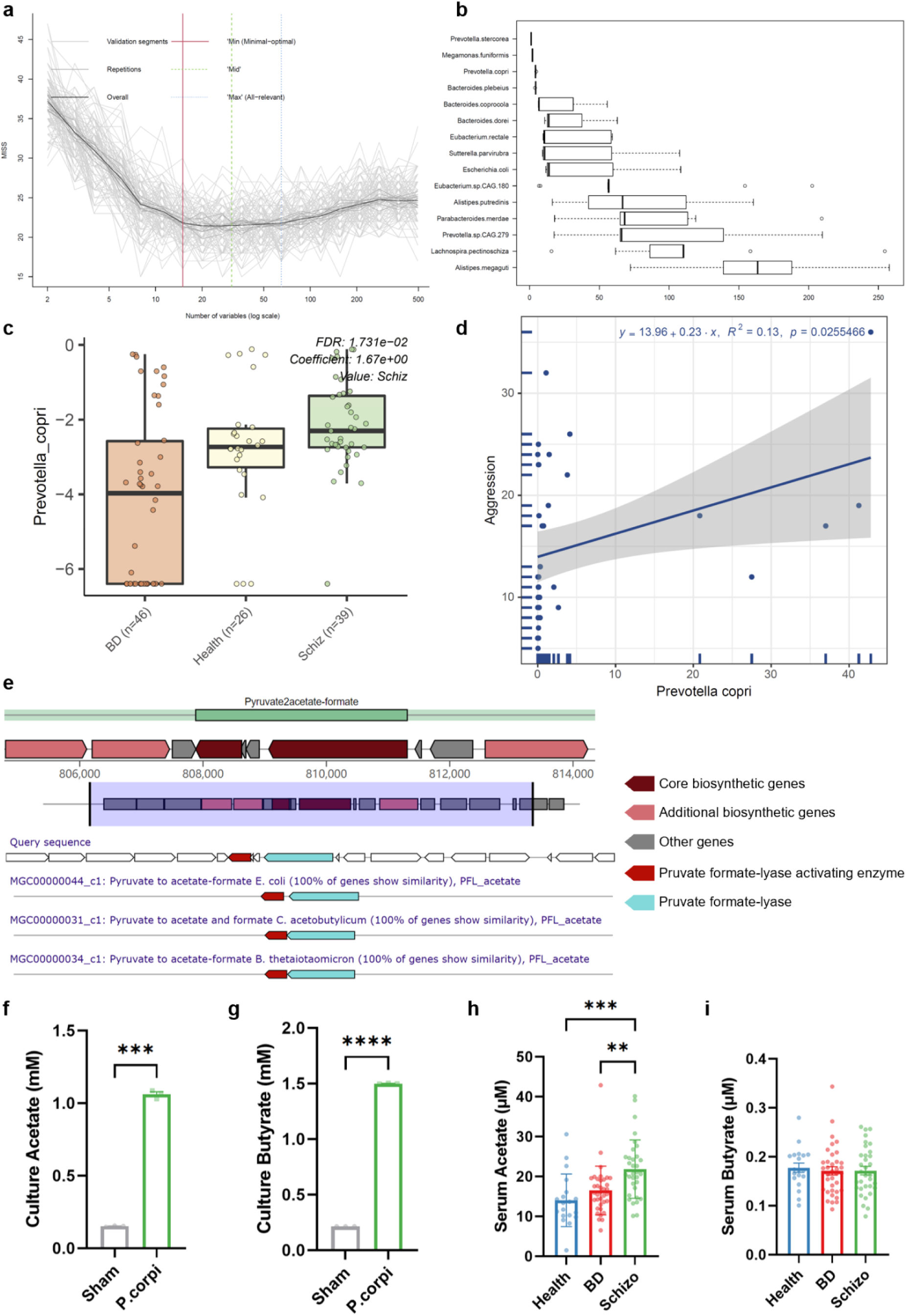
The acetate-producing *P. copri* is associated with schizophrenia. **a** Individual validation performance is indicated by light grey lines, while averaged validation curves for inner segments are indicated by darker grey lines. The random forest model with the minimal-optimal (Min) and all-relevant (Max) selections represent the outer limits of variable selections where least misclassifications occur in validation. **b** The Optimal modelling performance boxplot of the variables selected. Reproducible variables with low rank (lower is better) may be the strongest predictors. **c** Significant associations between relative abundance of *Prevotella copri* and schizophrenia at a threshold of q-value < 0.05. **d** The linear regression is significant at both sites of relative abundance of *P. copri* and PANSS hostility scores within schizophrenia patients at a threshold of *p* < 0.05. **e** MGCs annotation in the genome of *P. copri* type strain, predicting its acetate-producing ability, which is further proved by the quantification of acetate (**f**) and butyrate (**g**) after anaerobic inoculation (n = 3 per group). The serum concentrations of acetate (**h**) and butyrate (**i**) also support above results. Data are represented as mean ± SEM. ** *p* < 0.01, *** *p* < 0.001, **** *p* < 0.0001.

### Increased intestinal engraftment of *P. copri* promoted schizophrenia-like behavior and dysregulation of gut-brain serotonin metabolism in mice

To investigate potential *P. copri* involvement in schizophrenia-like behavior, we designed a mouse trial where intestinal microbes were depleted with a cocktail of antibiotics followed by two-week oral gavage of *P. copri* type strain DSM 18205 (Fig. 3a). The noncompetitive NMDA antagonist dizocilpine (MK801) has been commonly used to generate schizophrenia animal models, inducing the full spectrum of schizophrenia symptoms [28,29]. The treatment of antibiotics, MK801 or *P. copri* strain did not affect weight gain (Fig. 3b). In behavioral experiments, no significant differences in the response to background (70 dB) were observed between groups (Fig. 3c). When the pre-pulse intensity (PPI) was raised from 70 to 85, both blank and antibiotics groups maintained the same level of PPI, also suggesting that antibiotics treatment did not significantly affect the test. However, mice treated with either dizocilpine or *P. copri* strain showed significant PPI deficit at each higher intensity compared to the antibiotics group (Fig. 3d-f). The social preference index was significantly lower in MK801 group, while the *P. copri* strain failed to reduce sociability (Supplementary Figure 5). Based on the important role of the PFC in impaired cognition induced by MK801 [30], the Morris water maze (MWM) test was performed as an additional evaluation. Unexpectedly, the percent time spent in the target quadrant (Supplementary Figure 5) as well as the percent distance (Supplementary Figure 5) were not significantly affected by either MK801 or *P. copri* strain. Other behavioral experiments are detailed in the Supplementary Figure 5. The serum concentrations of acetate (Fig. 3g) and butyrate (Fig. 3h) were significantly increased in mice with *P. copri* strain, as expected. Similar trends were observed in the PFC without significant differences (Fig. 3i and j).

**Fig. 3.**
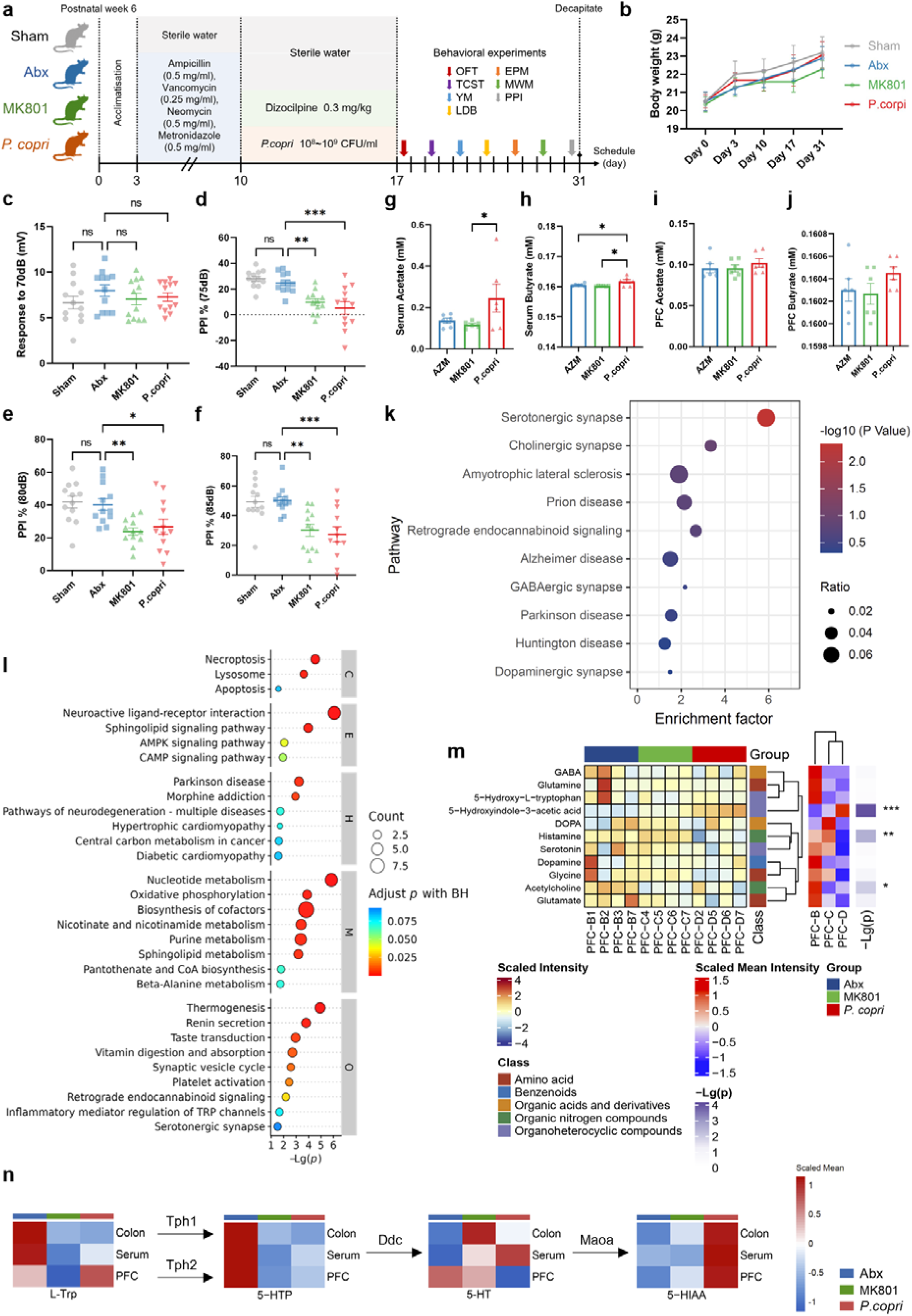
Multi-omics analysis based on induced schizophrenia-like behaviors after increasing engraftment of *P. copri* type strain. **a** Mice experimental design. **b** Body weight changes before behavioral experiments (n = 12 per group). **c** Similar response to background noise (70 dB) in the four groups. Both MK801 and *P. copri* type strain reduce the effect size of percent PPI impairments at 75 dB (**d**), 80 dB (**e**) and 85 dB (**f**) (n = 12 per group). **g-j** The quantification of acetate in serum (**g**) and PFC (**h**) as well as butyrate in serum (**i**) and PFC (**j**) (n = 6 per group). **k** Enrichment analysis and neurological selection of differentially expressed genes in PFC after *P. copri* engraftment (n = 4 per group). **l** Enrichment analysis of differential metabolites in PFC after *P. copri* engraftment (n = 4 per group). **m** The quantification of neurotransmitters in PFC and comparison among groups (n = 4 per group). **n** Scaled abundance of metabolites involved in the process of serotonin biosynthesis and degradation (n = 4 per group). Data are represented as mean ± SEM. * *p* < 0.05, ** *p* < 0.01, *** *p* < 0.001, ns depicts no significant difference.

Next, both transcriptome and metabolome analysis were performed to explore pathways affecting PFC. A total of 131 genes (49 upregulated and 82 downregulated after engraftment of *P. copri* strain) were differentially expressed (Supplementary Figure 6). Surprisingly, differentially expressed genes from the PFC were enriched in the serotonergic synapse (Fig. 3k), further corroborating the annotation of GBMs from clinical metagenome (Fig. 1f). The enrichment analysis of untargeted metabolome in the PFC also supported above results, to some extent, although these serotonin-related metabolic modules or pathways were non-targeted or statistically non-significant (Fig. 3l). Furthermore, the quantitation of neurotransmitters in the PFC was performed. Of these, the concentration of 5-hydroxyindolw-3-acetic acid (5-HIAA) was significantly increased after engraftment of *P. copri* strain, while histamine and acetylcholine were significantly decreased (Fig. 3m). The decreasing tendencies were observed in the PFC concentrations of 5-hydroxy-L-Trp (5-HTP) and serotonin (5-HT), although differences did not reach statistical significance (Fig. 3m). Notably, the changes of 5-HTP and 5-HT were not consistent in the colon, serum and PFC (Fig. 3n). The L-Trp catabolism was enhanced in mice colon after treatment with MK801 or *P. copri* strain, while more L-Trp was enriched in the PFC after *P. copri* gavage (Fig. 3n). As this process continued, the content of 5-HT was increased in the colon (Supplementary Table 6) and transported into the serum (Supplementary Table 7), finally degraded to 5-HIAA. However, the concentration of 5-HT was lower in the PFC of MK801 and *P. copri* group (Fig. 3m). It seemed that more 5-HT was degraded after increasing intestinal engraftment of *P. copri*, especially in the PFC (Supplementary Table 8).

### Acetate regulated serotonin biosynthesis, transportation and degradation in the colon and PFC

Another *in vivo* experiment was conducted to test whether the acetate could act independently (Fig. 4a). The treatment of MK801 or acetate did not significantly reduce weight gain (Fig. 4b). Consistent with expectations, the acetate added in drinking water for 4 weeks caused significant PPI deficit at all intensities (Fig. 4c). Unlike the *P. copri* strain, the acetate independently aggravated cognitive function impairment in mice, which was also consistent with the MK801 group (Fig. 4d and e). During the training trials of MWM, significantly differences in the escape latency were obtained at day 4-5 (Supplementary Figure 7). However, other behavioral experiments were little or no affected by acetate treatment (Supplementary Figure 7). To validate the above hypothesis of gut-brain serotonin metabolism, the protein expressions in colon (Fig. 4f) and PFC (Fig. 4g) were evaluated by Western blotting. For the biosynthesis of serotonin, the results demonstrated that significant increase in relative expressions of Tph1 (Fig. 4h) and Ddc (Fig. 4i) in the colon after treatment of MK801 or acetate. For the serotonin degradation, the protein expression of Maoa in the colon was significantly higher compared to blank controls (Fig. 4j). Expectedly, both MK801 and acetate enhanced the expression of Sert (Fig. 4k), which indirectly supported the distribution and changes of L-Trp and 5-HT in the colon and serum (Fig. 3n). Not only activated by acetate, the Ffar3 was also higher expressed in the colon after treatment of MK801 (Fig. 4l). The Ffar2 did not seem to be fully activated, despite being one of the main receptors for acetate (Fig. 4m). The free fatty acid receptor 3 (FFAR3, also termed as GPR41) in the brain has been shown to participate in the cognitive processes *in vivo*. The inactivation of FFAR3 prevented impaired cognitive function and pathological features associated with Alzheimer’s disease [31]. Regrettably, there is not enough evidence to interpret above phenomena associated with schizophrenia-like behaviors. The PFC protein expressions of Tph2 (Fig. 4n), Ddc (Fig. 4o), Maoa (Fig. 4p) and Sert (Fig. 4q) were consistently elevated after treatment of MK801 or acetate. Importantly, the increased synthesis and reuptake of 5-HT did not appear to be retained in the PFC, but was more degraded to 5-HIAA (Fig. 3n).

**Fig. 4.**
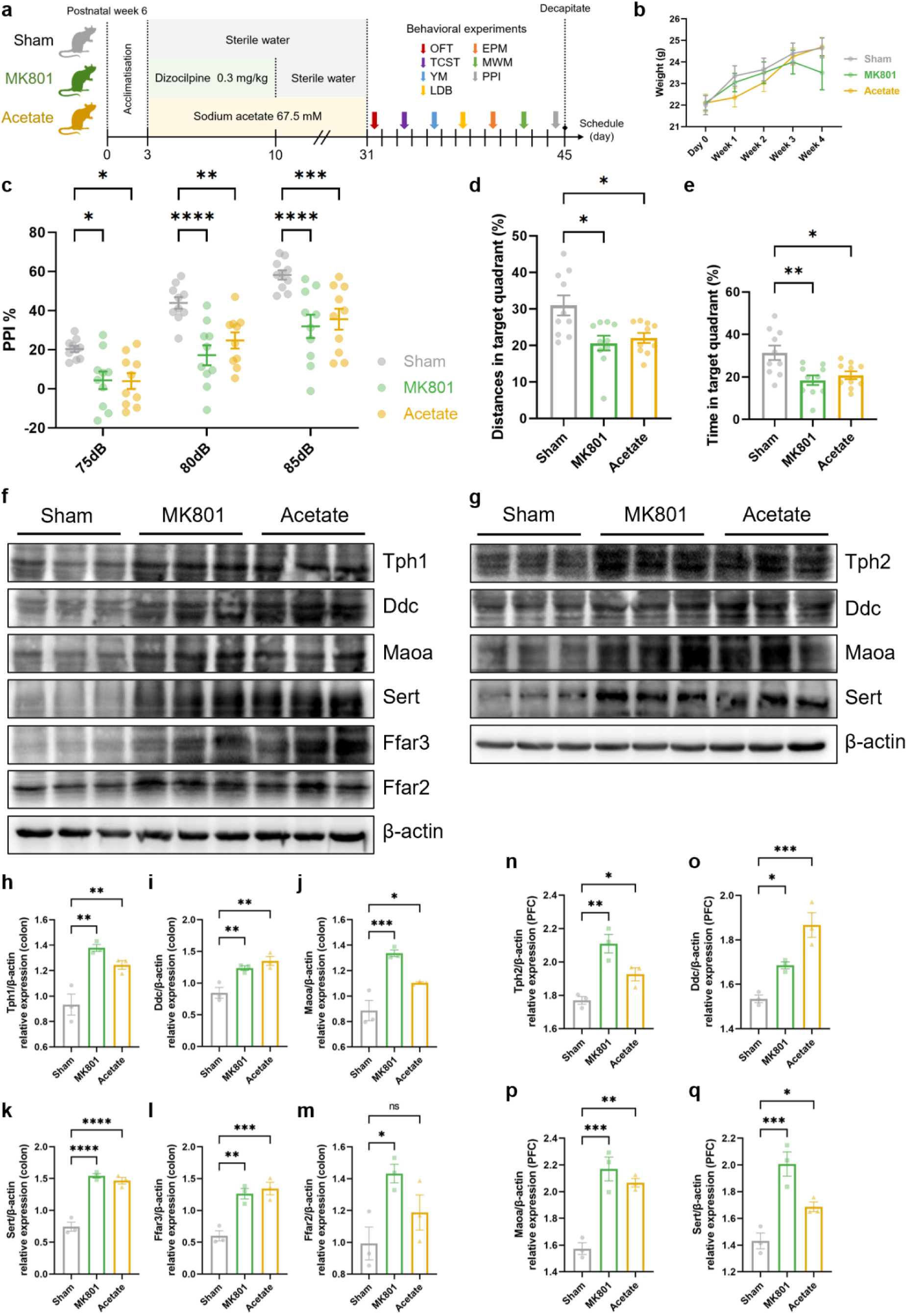
Increasing acetate intake regulated gut-brain serotonin metabolism. **a** Mice experimental design. **b** Body weight changes before behavioral experiments (n = 10 per group). **c** The effect size of percent PPI impairments at 75, 80 and 85 dB. The percent scores of distances (**d**) and time (**e**) spent in target quadrant of MWM test. **f-q** Western blot results of proteins expression in colon (**f**) and PFC (**g**). The quantification of Tph1 (**h**), Ddc (**i**), Maoa (**j**), Sert (**k**), Ffar3 (**l**) and Ffar2 (**m**) in the colon as well as Tph2 (**n**), Ddc (**o**), Maoa (**p**) and Sert (**q**) in the PFC were performed. Data are represented as mean ± SEM. * *p* < 0.05, ** *p* < 0.01, *** *p* < 0.001, **** *p* < 0.0001, ns depicts no significant difference.

## DISCUSSION

By using an integrated method to minimize the effects of clinical covariates, we identified the acetate-producing *P. copri* as one of the schizophrenia-specific metagenomic features, which was not reported before. The annotation of gut-brain functional modules based on clinical metagenome suggested a dysfunctional serotonin metabolism in schizophrenia rather than BD. These results were further proved in animal experiments through combining multi-omic approaches. The acetate was also shown to independently cause schizophrenia-related behaviors in mice and regulate their gut-brain serotonin biosynthesis, transportation and degradation. Since our analysis only cross-sectionally compared the gut microbiome of schizophrenia with BD, the changes of different strains of *P. copri* carrying similar functional genes in disease dynamics remain to be explored by future studies.

Previously, the family study was considered as a crucial test to determine diagnostic validity and to differentiate disease processes of schizophrenia and BD [32]. Nevertheless, a relative with schizophrenia or bipolar disorder was more likely than a relative without schizophrenia to have the disorder. Even though family history remained the strongest risk factor, co-segregation was also significant [33,34]. The Bipolar-Schizophrenia Continuum theory advocates a continuum encompassing unipolar disorder, BD, psychosis and schizophrenia, which is however more ambiguous [35]. Likewise, schizophrenia and BD shares some similarities in the dysfunction of neurotransmitter [36]. It is still difficult to diagnose and clearly distinguish even with official guidelines available, such as the Diagnostic and Statistical Manual of Mental Disorders (DSM) and the International Classification of Diseases (ICD). The gut microbiome have been demonstrated to play a crucial role in early-life neurogenerative processes [37], with perturbations inducing depressive-like behaviors in microbiota-necessary and host metabolism-mediated modes [38]. Recent studies have linked altered gut microbiome to neurological and neurodevelopmental disorders [39,40], as well as schizophrenia [18,41]. Depressive syndrome has been recognized as a risk factor for psychosis transition in vulnerable people [42]. The individuals who later developed schizophrenia had depression as the first premorbid signs [43], supporting the results of this study (Fig. 1c). Although the schizophrenia participants in this study were not first-onset, the variation explained by gut microbiome appeared to be persistent. Unfortunately, there is not enough evidence to explain this phenomenon and to determine which microbes may be involved in this process.

We identified the *P. copri* as an essential player in schizophrenia after excluding other species with any potential effects of clinical laboratory indicators and previous therapeutic time and modalities, although no significant differences in beta diversity were observed between schizophrenia and BD. It has been emphasized that the methodologies differ greatly and lack reproducibility across mental-disease studies [44]. Due to population differences, dietary patterns and medication, the metagenome-wide association study (MWAS) has been controversial for gut microbiome [20]. Current systematic reviews have showed that gut microbiome share some taxonomic commonalities between schizophrenia and BD [44,45]. Although only few studies were included and limited clues were provided, the genus-level *Prevotella* enriched in schizophrenia patients was indicated in the cross-disease meta-analysis [45]. An inclusion of more samples seems to achieve most convincing results of similarities and differences in taxonomic composition. Similarly, a higher level of *Prevotella* was highlighted in schizophrenia patients compared to healthy or normal controls. In contrast, patients with major depressive disorder had a lower level of *Prevotella* in the gut. The relative abundance of genus-level *Prevotella* appeared to be a schizophrenia-specific feature, although similar or opposite alternations were not reported in BD patients [44]. At the species level, our study firstly reported that the *P. copri* correlated closely with schizophrenia patients and could independently aggravate schizophrenia-like behaviors in mice. Given the high strain-level diversity and functional heterogeneity of the *P. copri* [26,27], more strains carrying same genes and acting similar functions may continue to be discovered. Although it is difficult to rank the importance of intestinal engraftment and actual functions, it is necessary to discover and verify functional modules contained in these strains.

Among functional modules annotated from clinical metagenome, we observed an increasing biosynthesis of serotonin (5-HT) in schizophrenia compared to BD. It has been proposed that subcortical 5-HT function increases during positive symptoms and prefrontal 5-HT function decreases during negative symptoms as the results of a cortical-subcortical imbalance in schizophrenia patients [46]. Similarly, a low level of 5-HT has been detected in the PFC, which partially mediates the negative symptoms of schizophrenia [47]. Of note, the inconsistency of 5-HT levels in colon, serum and PFC was observed after treatment of *P. copri* type strain in our study (Fig. 3n). The correlation between 5-HT and Trp was significantly positive in healthy individuals, while this correlation was lost in both first-onset and post-treatment schizophrenia patients [48]. Previous evidences have supported a higher degradation of Trp in schizophrenia patients [49,50]. The CSF levels of 5-HIAA showed a significant increase in schizophrenia patients and returned to normal after clinical recovery [51]. Likewise, the significantly positive correlation between 5-HIAA and Trp in healthy individuals was also lost in both first-onset and post-treatment schizophrenia patients [48]. Based on the fact that over 90% of 5-HT in the host originates from the gut, potential moderation effects of engrafted bacterial strains were too strong to ignore in the biosynthesis, degradation and transportation of 5-HT.

We found that the acetate independently caused schizophrenia-like behaviors without increasing intestinal engraftment of *P. copri* type strain. A previous study has been demonstrated that the *P. copri* can catabolize dietary fibers and produce SCFAs, which protect mucosal barrier and lower inflammation potential [52]. Despite this, we still confirmed the acetate-producing ability of P. copri type strain *in vitro*. By contrast, other studies linked a higher abundance of *P. copri* with new-onset rheumatoid arthritis [53,54] or impaired glucose tolerance [55]. Recent research suggested that the functional module of acetate degradation is significantly decreased in young schizophrenia patients, although no further validation was performed [56]. In this study, the acetate was firstly proved to independently promote schizophrenia-like behaviors, even similar to those produced by MK801 as a classical inducer of schizophrenia models. A four-week continued treatment of acetate significantly increased the expression of TpH1 (Fig. 4h) and TpH2 (Fig. 4n) *in vivo*. The TpH1 is significant associated with schizophrenia, while the minor role of TpH2 in schizophrenia has been reported in a genome-wide association study [57]. In addition, the single-nucleotide polymorphisms of aromatic-L-amino-acid decarboxylase (DDC) were identified as potential predictors of treatment-resistant schizophrenia patients [58]. A relatively higher level of 5-HT was observed in colon and serum after the treatment of *P. copri* (Fig. 3n), which was also indirectly supported by the results of acetate-mediated higher expressions of Ddc in both colon (Fig. 4i) and PFC (Fig. 4o). Somewhat to our puzzlement, the acetate not only enhanced the biosynthesis of 5-HT but also facilitated the degradation (Fig. 4j and 4p). Notably, gene polymorphisms of monoamine oxidase A (MAOA) and serotonin transporter (SERT) were demonstrated the major roles in the depressive symptoms of schizophrenia [59], supporting the results of variable explanation of clinical metagenome to PNASS scores in schizophrenia patients (Fig. 1c). The SERT inhibitors have been commonly used to improve depressive behaviors by enhancing extracellular 5-HT levels in the brain. However, the treatment-resistant patients appeared to respond to higher levels of 5-HT derived from 5-HTP than those produced by SERT inhibitors. The administration of 5-HTP in slow-release delivery safely elevated brain 5-HT beyond the therapeutic effects of SERT inhibitors [60]. In part, the gut-brain inconsistency in the metabolic progress from Trp to 5-HIAA was illustrated.

Our study provides unique evidence that the acetate-producing species *P. copri* selected from clinical metagenome can aggravate schizophrenia-like behavior in mice. To be accurate, potential effects of clinical variables were evaluated as fully as possible and the *P. copri* was identified as one of the schizophrenia-specific features at taxonomic level. The dysfunctional serotonin metabolism was observed in schizophrenia rather than BD, which was schizophrenia-specific at functional level. These metagenomic features were further proved by using multi-omic approaches *in vivo* and *in vitro*, representing a blueprint of microbiome-colon-PFC communications in schizophrenia. Together, these observations may open up new perspectives for schizophrenia differential diagnosis and post-treatment surveillance.

### Limitations of the study

This study merits consideration in several aspects. First, the relatively complete metagenome-assembled genomes (MAGs) may be more suitable to be used as microbial biomarkers for further explorations of mechanism and clinical application. However, it was failed to be generated high-quality MAGs of different strains of *P. copri* through assembly and binning methods in this study. Second, a longitudinal study design seems to be needed to increase the accuracy of taxonomic classification and functional annotation, although only machine-learning selection based on a small sample size with *in vivo* validation may not be the first choice for screening out gut microbial biomarkers in complex mental diseases. Third, different dosing responses to the acetate have not been fully evaluated in this study, which is necessary to further investigate in the future.

## METHODS

### Cohort recruitment

The healthy controls and patients with schizophrenia and bipolar disorder were recruited at the Guangzhou Huiai Hospital, from October 2017 to December 2018. According to the Diagnostic and Statistical Manual of Mental Disorders fourth Edition (DSM-IV), diagnoses were established. For schizophrenia, patients with acutely relapsed schizophrenia and first-episode schizophrenia were recruited into this study. An evaluation of clinical psychopathological symptoms was conducted by the Positive and Negative Syndrome Scale (PANSS), which was further classified into six sub-scales (anergia: N1 + N2 + G7 + G10; thought disturbance: P2 + P3 + P5 + G9; activation: P4 + G4 + G5; paranoid: P6 + P7 + G8; depression: G1 + G2 + G3 + G6; aggression: P4 + P7 + G6 + S1 + S2 + S3) as previously reported [61,62]. For bipolar disorder, the Bech-Rafaelsen Mania Scale (BRMS) and 24-item Hamilton Depression Rating Scale (HAMD-24) were used in the initial diagnostic assessment. Exclusion criteria were that the patient had received antipsychotic treatment (drug or non-drug) from other medical institutions or self-administered within three months before the current hospitalization. Antipsychotic treatments received by the patients before the three-month treatment gap period were recorded based on previous medical records to minimize potential impact of this variable. Any participants were excluded if: 1) they had taken antibiotics (oral or injected) in the previous three months, or 2) they had a suspected or diagnosed bacterial, fungal, or viral infection, constipation or diarrhea, or 3) their stool was abnormal (according to the Bristol Stool Scale).

### Mice

The specific pathogen-free (SPF) C57BL/6 mice (6 weeks old, 20 ± 2g) were purchased from the Guangdong Medical Laboratory Animal Center (Guangzhou, China). After 72 hours of acclimatization to animal laboratory conditions, mice were randomly assigned into different groups according to study designs. In the first study, the antibiotic cocktail consisting of ampicillin (0.5 mg/ml), metronidazole (0.5 mg/ml), neomycin (0.5 mg/ml), and vancomycin (0.25 mg/ml) was administered to mice for 7 days, which was modified base on previous protocol [63]. A relatively equal number of *P. copri* strain (same passage number and same experimental batch, OD600 = 0.9 ± 0.05, about 8 × 10^8^ CFU/ml) was prepared daily and orally gavaged for 2 weeks. In the second study, the sodium acetate was dissolved in sterile water to achieve a concentration of 67.5 mmol/L according to the previous study [64]. The drinking water containing sodium acetate was replaced every 2 days for 4 weeks. The dizocilpine (MK801) (0.3 mg/kg) was administered after mice had been habituated for 30 minutes. The intraperitoneal injection was performed daily for 7 days.

### Samples collection

For the clinical samples, blood and stool were collected on the day when two psychiatrists conducted independent assessments. The blood samples were centrifuged at 3,000 g for 15 minutes after remaining still for 0.5 hours in room temperature. The stool samples were collected by the patient or with the help of family members using three sample tubes and transferred to −80 within 30 minutes. For the mouse samples, preparation of serum was consistent with above method. The colon and PFC tissues were collected and quickly quenched in liquid nitrogen. The frozen tissue samples were lyophilized and the dry weight was determined for subsequent experiments. All samples were aliquoted before transferring to −80 for storage to avoid repeated freeze-thaw.

### Metagenome library construction and sequencing

The extractions of DNA from 0.2 g of stool were performed according to the manufacturer’s standard procedure with the PowerSoil DNA Isolation Kit (MoBio Laboratories, Carlsbad, CA, USA). Extracted DNA was stored at −80 until use. All DNA samples were quantitated using the Qubit dsDNA HS Assay Kit (Invitrogen, Carlsbad, CA) based on Qubit 2.0 (Thermo Fisher Scientific, Waltham, MA, USA), then normalized to a final concentration of 50 pg/mL. The paired-end libraries were prepared and sequenced on the Illumina Miseq platform (Illumina Inc., San Diego, CA, USA).

### Taxonomic and functional metagenome profiling

The metagenomic sequencing reads were quality controlled by removing adaptor sequences using fastqc v0.11.9, followed by trimming low quality reads and removing human sequences using KneadData v 0.10.0. Taxonomic profiles were generated by MetaPhlAn2 v2.7.7 (31 May 2018) at the species level [65]. Functional profiles were generated using HUMAnN2 v2.8.1 [66]. The annotation of gut-brain modules (GBMs) and gut metabolic modules (GMMs) was performed using omixer-rpmR v0.3.2 [67]. The primary metabolic gene clusters (MGCs) were identified by gutSMASH v1.0 (October 2020) [68].

### Transcriptome library construction and sequencing

The extractions of RNA from 20 mg of PFC tissues were performed according to the manufacturer’s standard procedure. Extracted RNA samples were evaluated using the NanoDrop (NanoDrop Technologies, Wilmington, NC, USA), followed by Qubit 2.0 and Agilent 2100 (Agilent Technologies, Santa Clara, CA, USA) for absolute quantification. Quantitated RNA samples were then processed for library construction. The purification of cDNA was performed using AMPure XP beads (Beckman Coulter, Brea, CA, USA). For ensuring the quality of libraries, Qubit 2.0 and Agilent 2100 were also used to measure cDNA concentration and insert size. The paired-end sequencing was carried out by the Biomarker Technologies (Beijing Biomarker Technologies Co., LTD., Beijing, China).

### Transcriptomic analysis

The transcriptomic sequencing reads were quality controlled using fastqc v0.11.9. Reference genome (mus_musculus.GRCm38_release95) for the analysis was predefined. HISAT2 v2.2.1 [69] was used for mapping reads, followed by assembling the mapped reads using StringTie v2.2.0 [70], which was also used to discover potential novel transcripts based on reference genome. The transcripts with coding peptides length less than 50 amino acids or containing only one exon were excluded. The maximum flow algorithm of StringTie v2.2.0 was used to measure the expression level of a gene or transcript using FPKM (Fragments Per Kilobase of transcript per Million fragments mapped). The annotation of differentially expressed genes was on the basis of KEGG (Kyoto Encyclopedia of Genes and Genomes) database. A total of 10 pathways were selected according to the key words (nervous and neuro) among 43 categories at the second level of KEGG annotation and ranked based on statistical values.

### Metabolomic sample preparation

Prior to extraction of metabolites, lyophilized colon and PFC tissues (20 mg wet weight) were ground for one minute in a Grinding Mill (Servicebio Technology Co., Ltd., Wuhan, China) at 65 Hz with 5 mm tungsten beads. Precooled mixtures of methanol, acetonitrile, and water (v/v/v, 2:2:1) were used to extract metabolites from tissues. For serum, an equal volume of methanol acetonitrile (v/v, 1:1) was mixed with samples by vortexing for 5 minutes. The mixtures were under ultrasonic shaking for one hour and kept at −20 for another hour, followed by centrifuging at 16,000 g for 20 minutes at 4. The supernatants were moved into another 2 mL tube and concentrated to dryness in a vacuum centrifuge concentrator (Eppendorf, Hamburg, Germany).

### Metabolome profiling

The prepared samples were separated with liquid chromatography (LC) using a ACQUITY UPLC HSS T3 column (2.1×100 mm, 1.8 μm) (Waters, Milford, MA, USA). The mobile phases were 0.1% formic acid (A) and 100% acetonitrile (B). The gradient was 0% buffer B for 2 min and linearly increased to 48% in 4 min, then up to 100% in 4 min and maintained for 2 min, finally decreased to 0% buffer B in 0.1 min, followed by a 3 min re-equilibration period. The flow rate was set as 0.3 mL/min. For data acquisition, electrospray ionization (ESI) with positive and negative modes was separately utilized based on a Nexera X2 LC-30AD UHPLC (Shimadzu, Kyoto, Japan) coupled with Q-Exactive Plus Orbitrap hybrid mass spectrometer (Thermo Fisher Scientific, Waltham, MA, USA). Main parameters were set as follows: spray voltage: 3.8kV (positive) and 3.2kV (negative), capillary temperature: 320, sheath gas (nitrogen) flow: 30 arb (arbitrary units), aux gas flow: 5 arb, probe heater temperature: 350, S-Lens RF (radio frequency) level: 50. For the full MS scan (from 70 to 1050 Da), a resolution of 70,000 was acquired at m/z 200. For the MS/MS scan, a resolution of 17,500 was acquired at m/z 200. Injections for full MS and MS/MS were limited to 100 ms and 50 ms, respectively. As for MS2, the isolation window was set as 2 m/z and the normalized collision energies as 20, 30 and 40. After acquiring raw data, the peak alignment, retention time correction and peak extraction were performed using MS-DIAL v4.24 as previously reported [71]. The msp databases (VS17) from authentic standards, human metabolome database (HMDB) and self-built metabolite standard library (Bioprofile Biotechnology Co., Ltd., Shanghai, China) were selected for identification. The variables that had more than 50% of nonzero measurements in at least one group were retained in the extract-ions.

### Quantification of short chain fatty acids

Serum samples without any treatment and lyophilized PFC tissues redissolved in methanol were transferred to precooled sampling vials on ice and sealed as soon as possible. The quantification of short chain fatty acids was performed based on the Agilent 7697A headspace sampler equipped with an Agilent 7000D triple quadrupole mass spectrometer (Agilent Technologies, Santa Clara, CA, USA). The separation of short chain fatty acids was performed using a DB-WAX capillary column (30 m × 0.25 mm × 0.20 μm) (Agilent Technologies, Santa Clara, CA, USA). Each standard curve contained five different points at least (R2 ≥ 0.99).

### Quantification of neurotransmitters

Weighed dry samples were resuspended in the precooled mixture of an equal volume of methanol water (v/v, 1:1). After centrifuging at 20,000 g for 15 minutes at 4, the supernatants were moved into dark glass vials. The prepared samples were separated using a ACQUITY UPLC BEH C18 column (2.1×100 mm, 1.7 μm) (Waters, Milford, MA, USA). The mobile phases and flow rate were consistent with the above. The gradient was maintained at 2% of buffer B for 4 min, then increased to 80% from 4 to 6 min and to 90% from 6 to 8 min, then maintained at 90% for 2 min, finally decreased to 2% of buffer B in 1 min. The quantifications of 17 neurotransmitters were performed in multi-reaction monitoring (MRM) mode based on a Nexera X2 LC-30AD UHPLC coupled with QTRAP 5500 LC-MS/MS system (AB SCIEX, Framingham, MA, USA). Main parameters were set as follows: ion spray voltage floating: 5.5kV (positive) and −4.5kV (negative), source temperature: 550, ion source gas 1: 40psi,

ion source gas 2: 50psi, curtain gas (CUR): 35psi. The mass spectrometry detection was completed by the Bioprofile Biotechnology (Bioprofile Biotechnology Co., Ltd., Shanghai, China). A total of 11 neurotransmitters were quantified using external standard method (6 of 17 were not detected or below limit of detection). Each standard curve contained six different points (R2 ≥ 0.99).

### Anaerobic culture of bacterial strains

The type strain *Prevotella copri* DSM 18205 was purchased from DSMZ-German Collection of Microorganisms and Cell Cultures GmbH (Braunschweig, Germany). After incubating with Schaedler Broth (DSMZ Medium 1669) under anaerobic condition for 72 hours, bacteria strains were collected at mid-logarithmic growth phase (OD600 in the range of 0.8 to 1.0) and kept frozen at −80 in 20 % glycerol not more than one month. The thawed samples were centrifugated at 3000 g for 10 minutes at 4 and resuspended in sterile PBS before oral gavage. Another three independent batches with blank controls were centrifugated at 10,000 g for 20 minutes at 4, followed by filtering at 0.22 μm to remove bacterial cells and debris before quantification of short chain fatty acids.

### Behavioral experiments

The behavioral experiments were conducted between 9:00 AM and 6:00 PM for a continuous period. At least 10 mice per group were used for each test according to the previous study [18]. All of tests and outcome assessments were conducted by two independent investigators blinded to group allocation. The mice were habituated in the behavioral experiments room for one hour before testing. In order to minimize the influence of prior test history, the next test was performed in sequence from the least to the most invasive with a break of 48 hours at least. The raw data was accessed using the SMART v3.0.03 video-tracking system (Panlab, Barcelona, Spain) or recorded by manual observation. The behavioral experiments were carried out in the following order: open field test (OFT), three-chamber social test (TCST), Y-maze (YM) test, light-dark box (LDB) test, elevated plus maze (EPM), Morris water maze (MWM) and pre-pulse inhibition (PPI) test.

#### Open-field test

In an open-field arena (45 × 45× 45 cm), mice were allowed to explore freely for 30 minutes, and their distance travelled and time spent in the central zone were accessed. Any rearing activity and jumping were manually observed and recorded.

#### Three-chamber sociability test

The three chambers (60 x 40 x 23 cm) were separated by dividing chamber walls, allowing mice to access each chamber separately. It was allowed to explore freely for 10 minutes in the center chamber during the habituation phase without access to the side chambers. Afterwards, one of the side chambers was placed with a conspecific unfamiliar to the group (stranger), which either matched the conspecific’s sex or strain. A similar inverted wire cup containing a novel object was positioned in the nonsocial side chamber. For the sociability phase, the test mouse explored the three chambers in free movement for 10 minutes while a video camera recorded distances traveled and time spent in each chamber.

#### Light-dark box test

The light-dark box consisted of two chambers (light and dark) with an entrance that enabled mice to move between them. In the dark box, mice were each introduced individually and allowed to move around freely for 10 minutes. Investigators measured how long they spent in the light chamber and how often they switched between the two.

#### Y-maze

The Y-maze test was conducted by placing mice at the end of an arm with the brightness and allowing them to move freely through the maze for 7 minutes. The alternation was defined as a mouse successively entering all three arms without repetition. As a result, the percent alternation was calculated as follows: [number of alternations]/[(total number of arm entries - 2) × 100 %].

#### Elevated plus maze

In a plus-shaped maze with two open and two closed arms (30 cm long, 10 cm wide, 20 cm walls, and 1 m above the ground), mice were placed in the central square and the time spent in each arm was manually recorded with video-tracking system. In order to enter an arm, a mouse had to use all four paws. Hence, it was determined how much time the animal spent in open arms and the ratio between open/closed arms time could be used as anxiety measures, since anti-anxiolytic drugs decrease these quantities.

#### Pre-pulse inhibition test

First, five minutes of white-noise background (70 dB) was used to habituated the mice to the chamber. There were 80 trials in each test: 6 null trials, 68 prepulse-pulse trials and 6 pulse-alone trials. Intertrial intervals ranged from 10 to 20 seconds on average. A 120-dB stimulus burst of 40 ms was used in each null trial. During prepulse-pulse trials, 7 types of modes were presented randomly: a 40-ms prepulse stimulus that was 75, 80, or 85 dB followed by a 120-dB stimulus 100 ms after the prepulse stimulus, a 40-ms burst of a 120-dB stimulus. The pulse-alone trials were conducted using the same protocol as null trials at the end of each test. The PPI responses were calculated as follows: [1-(prepulse trials/pulse-alone trials)] × 100%.

#### Morris water maze

An encased platform was hidden behind a circular plastic pool filled with water that was painted white with nontoxic paint. There were four equal quadrants in the pool, with the platform placed at the center of each quadrant. After training the animals for four days (six times a day), a probe test was conducted 24 hours later. The probe test involved removing the platform and measuring how much time was spent in each quadrant. A daily variation in starting positions and timing of reaching the platform was recorded.

### Western blot

The information for antibodies is as follows: Tph1 (D10C10) Rabbit mAb (#12339, Cell Signaling Technology), Tph2 (D3E5I) XP® Rabbit mAb (#51124, Cell Signaling Technology), Ddc (D6N8N) Rabbit mAb (#13561, Cell Signaling Technology), Maoa (E3L3B) Rabbit mAb (#75330, Cell Signaling Technology), anti-serotonin transporter antibody (EPR26259-61, abcam), FFAR3 antibody (CSB-PA008939, Cusabio Technology), FFAR2 Polyclonal antibody (19952-1-AP, Proteintech Group), β-Actin (13E5) Rabbit mAb (#4970, Cell Signaling Technology). The western blot was conducted in reference to previous report [72]. The quantification of immunoblots was performed using a ChemiDoc MP Imaging System (Bio-Rad, Hercules, CA, USA). After a normalization to β-actin, the band intensities of target protein were quantified using ImageLab v4.1.

### Statistics analysis

The results on behavioral experiments and quantification of acetate and butyrate of participants and mice were compared among groups using one-way ANOVA followed by the Dunnett correction for multiple comparisons or Kruskal-Wallis test followed by the Dunn’s correction. The changes of weight were compared among groups and time using two-way ANOVA followed by the Fisher’s LSD. The quantification of acetate and butyrate from bacterial culture was compared using unpaired t test with Welch’s correction. The results on relative expression of proteins were compared using one-way ANOVA followed by the Fisher’s LSD. The statistical calculations above were performed using Graphpad Prism v9.0.0. Other statistical analysis and visualization were performed using R version 4.1.3. The Bray-Curtis distance with normalization was used to generate PCoA ordination plot, which was visualized by ggplot2 package v3.3.6 and ggthemes package v4.2.4. The marginal histograms on both sides were added using ggExtra package v0.10.0. The PERMANOVA analysis was conducted with the adonis2 function from vegan package v2.6-4. The selection of significant differences in species-level taxonomy and identification of potential covariates were performed by MaAsLin2 package v1.6.0 [73] with a threshold of q-value < 0.05. The selection of significant differences in GBMs and GMMs and exclusion of the species with any potential effects of different antipsychotic treatments before the three-month gap period were conducted by lefser package v1.12.1 using the Kruskal-Wallis test, Wilcoxon-Rank Sum test and linear discriminant analysis (LDA). A random forest model was used as multivariate method to identify significant differences in taxonomic abundance at species level based on MUVR package v0.0.975 [74]. The ratio of variables to include in subsequent inner loop iteration and the number of repetitions of double cross-validation were set as default parameters (varRatio = 0.75, nRep = 5), and the number of external cross-validation (nOuter) was set as 10. The minimum-response variables in the model were selected and ranked according to the accuracy of their probability of classified prediction. The linear regression analysis and visualization were performed by the Hiplot [75]. The differential expression analysis was processed by edgeR v 3.36.0 [76] with thresholds of fold change (FC) ≥ 1.5 and false discovery rate (FDR) adjusted p-value (q-value) < 0.05. The multivariate analysis of metabolome was performed using SIMCA v14.1 and MetaboAnalyst v5.0 [77] with thresholds of |log2(FC)|>1 and VIP>1 with cvSE of VIP less than the VIP value. The KEGG pathway enrichment analysis was performed by clusterProfiler v4.0 [78] and significance of pathway was determined by enrichment factors and Fisher’s exact test.

## Data Availability

All data produced in the present study are available upon reasonable request to the authors.

## Declarations

### Ethics approval and consent to participate

This study was approved by the Medical Ethics Committee of Guangzhou Huiai Hospital and registered with the Chinese Clinical Trial Registry (ChiCTR-ROC-17012985). It was followed throughout this study all institutional requirements regarding the ethical use of human information and samples. The signed informed consents were obtained from each participant or his/her legal guardian before enrolling. Mice experiments were approved by the Animal Ethics Committee of Guangzhou University of Chinese Medicine (Approval No. 20220617001) before the beginning of *in vivo* experiments.

### Availability of data and materials

All data produced in the present study are available upon reasonable request to the authors.

### Consent for publication

Not applicable.

### Authors’ contributions

S.J. drafted the manuscript and performed metagenome and metabolome analysis. M.S. assisted with sequencing and data analysis. Q.H. and B.P. applied animal ethics and performed animal experiments. S.W. recruited subjects and interpretated clinical assessment. Y.L. and M.W. performed western blot and assisted with animal experiments. X.Z. and Y.Y. contributed to collecting and processing samples. L.D. contributed to interpretating metabolome data. D.L. performed anaerobic culture. J.C., A.Z. and J.Z. assisted with animal experiments. Z.J., C.W. and X.C. assisted with collection of clinical samples. J.C. applied clinical ethics. L.Y. managed clinical data and assisted with application of clinical ethics. T.V. contributed to revising the manuscript and assisted with analytical methods. X.Z. conceived the study and assisted with revision of manuscript. All data were generated in-house and no paper mill was used. All authors agree to be accountable for all aspects of work ensuring integrity and accuracy.

## Supplemental figures

**Fig. S1.**
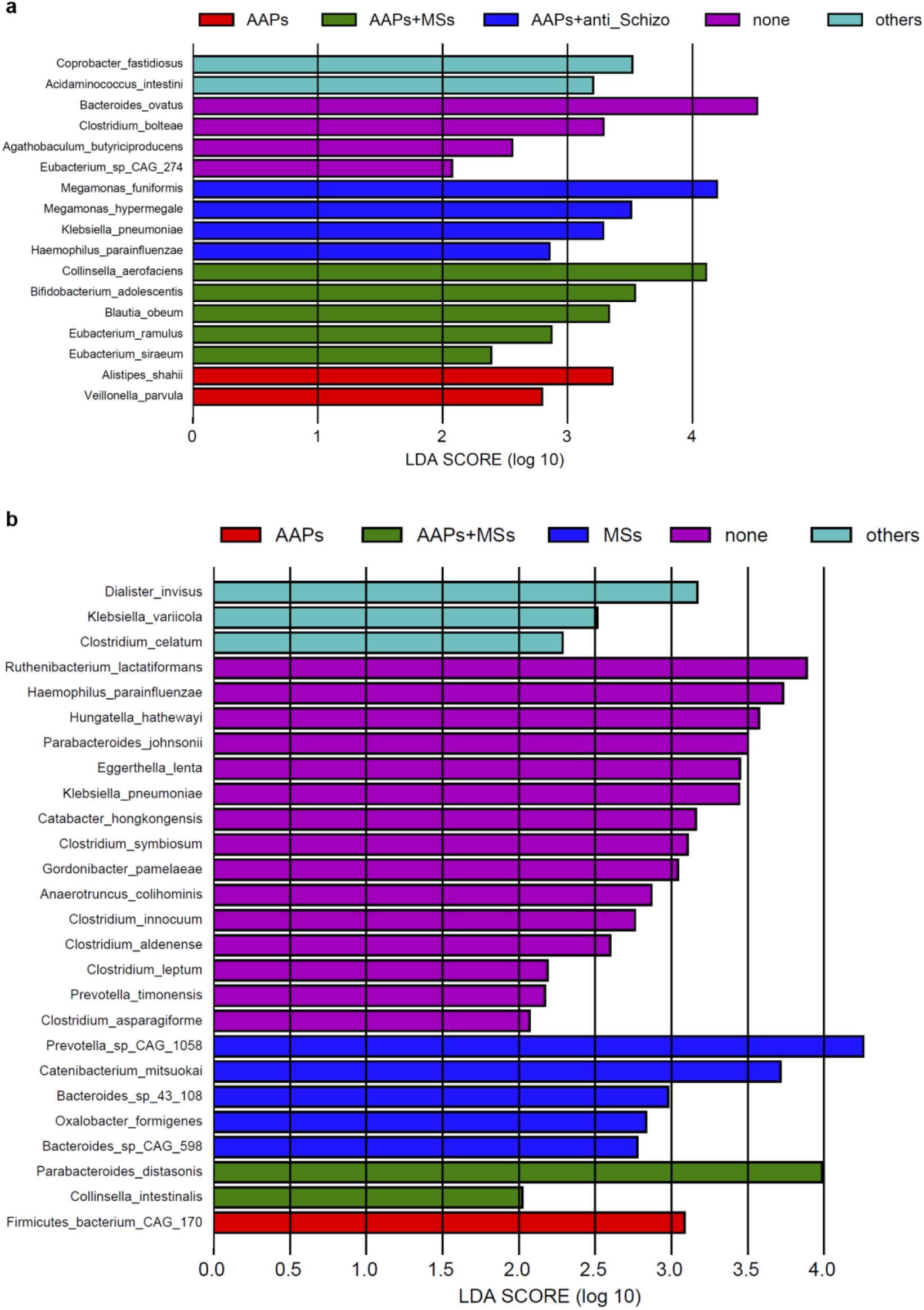
LEfSe analysis of potential effects of previous therapeutic modalities on gut microbiome in schizophrenia (**a**) and bipolar disorder (**b**) patients. Significances obtained by LEfSe analysis at p-value < 0.05 and LDA (log_10_) ≧ 2. (AAPs: atypical antipsychotics, MSs: mood stabilizers.)

**Fig. S2.**
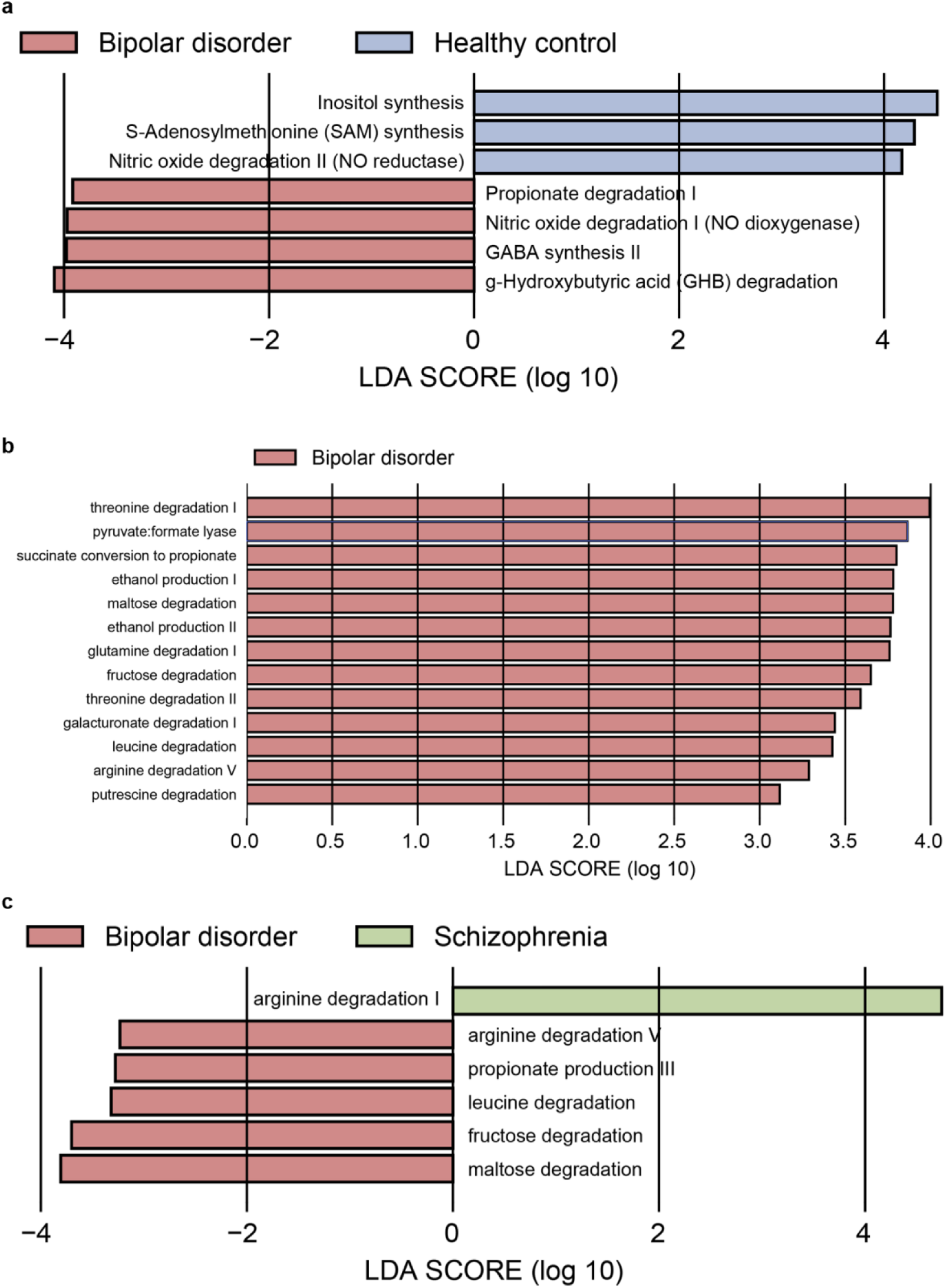
Group-wise comparisons of functional modules (GBMs: **a**, GMMs: **b**, **c**).

**Fig. S3.**
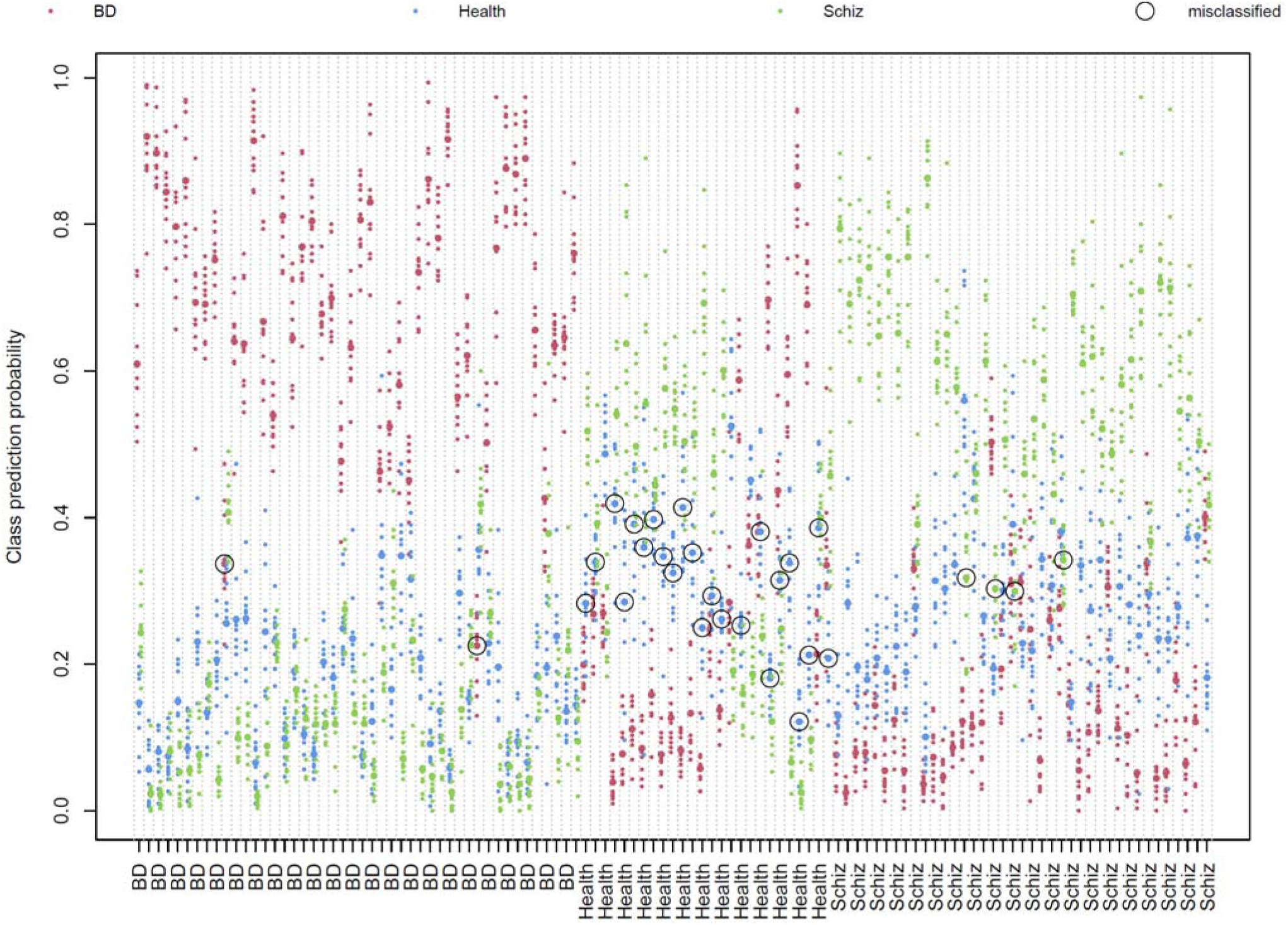
Class prediction probabilities of validation set using selected 15 species based on the optimal modelling performance of training set.

**Fig. S4.**
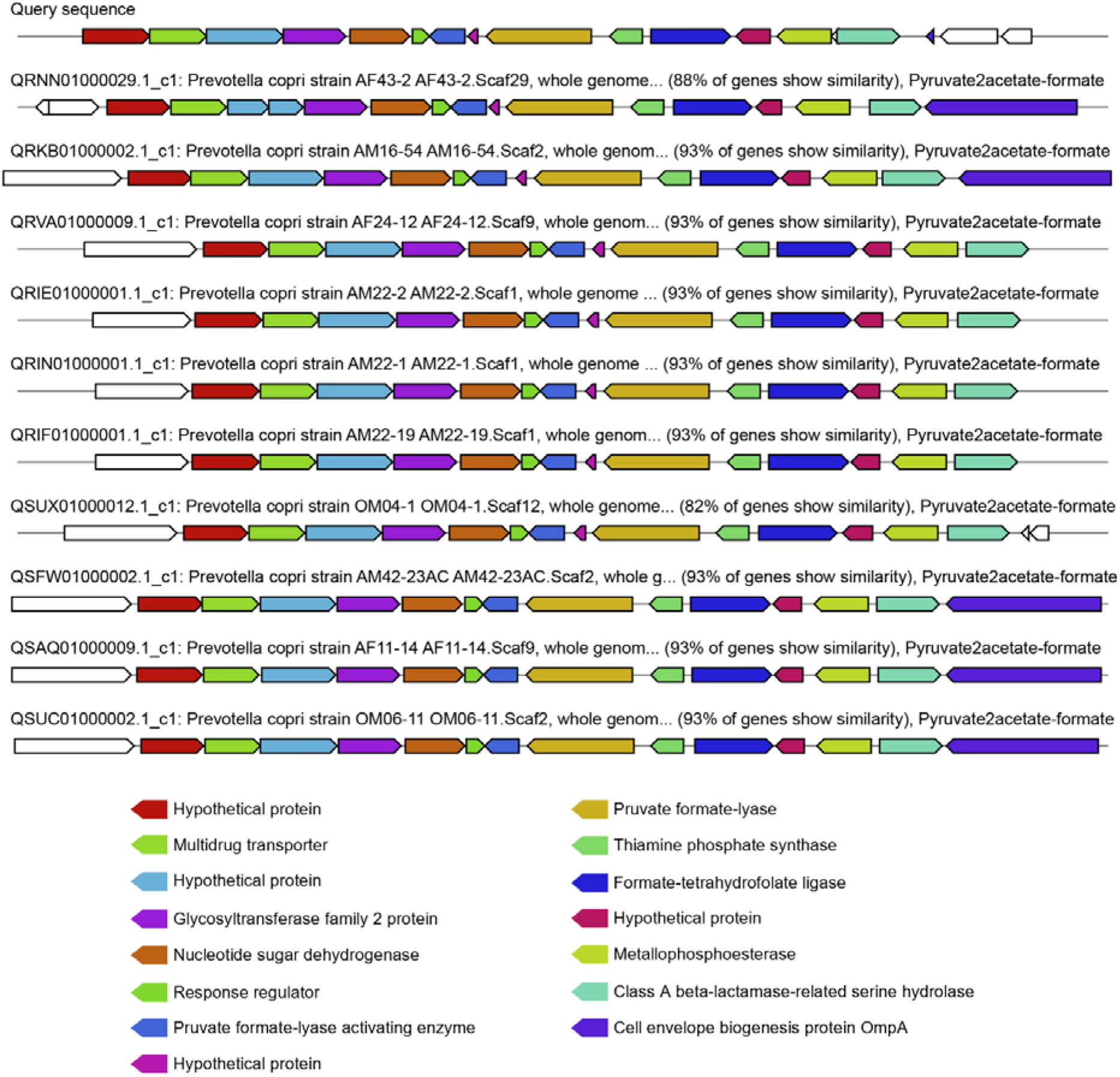
The MGC with high nucleotide similarity between queried sequences (GCA_000157935.1) and different strains of *P. copri*.

**Fig. S5.**
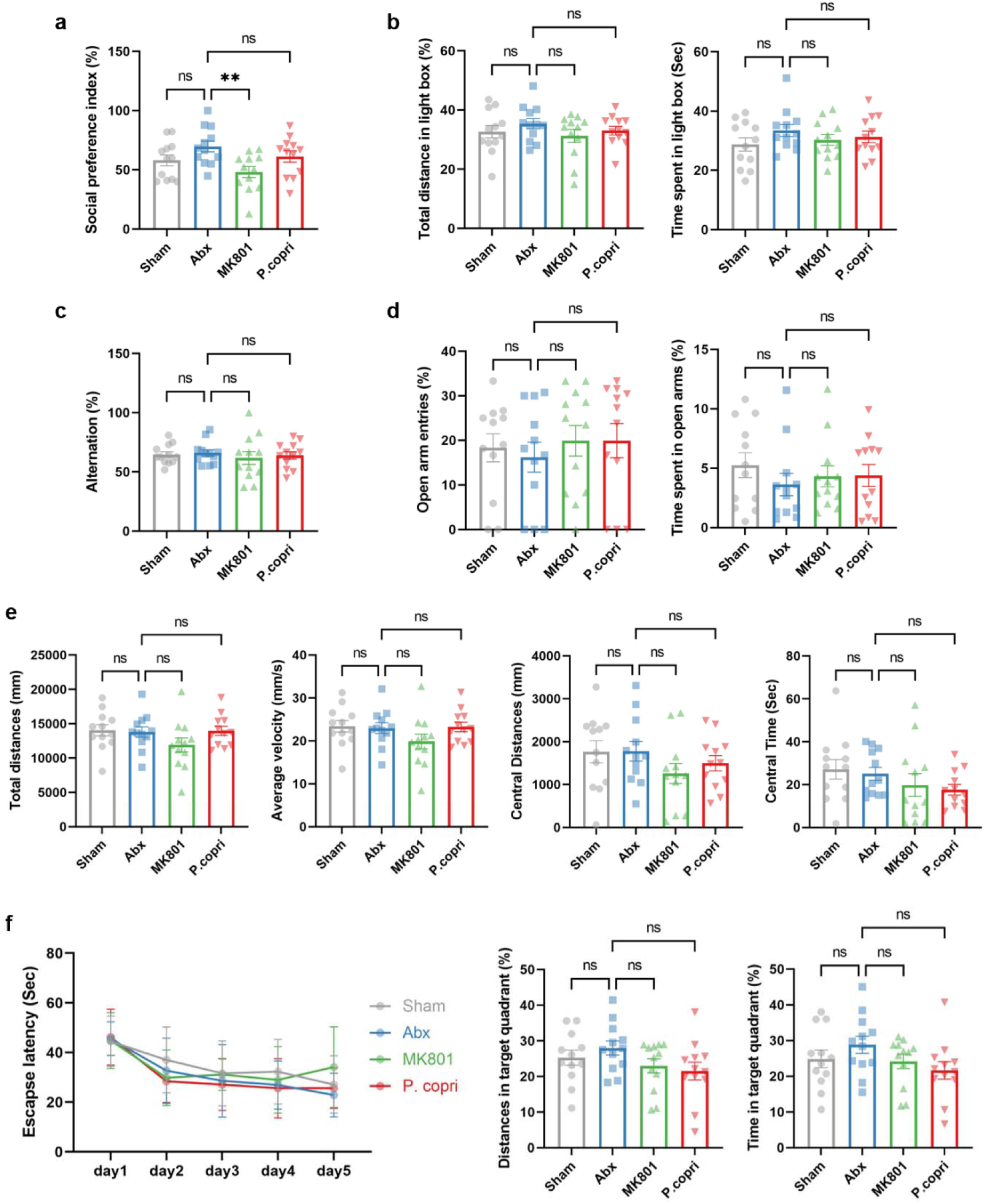
Behavioral experiments after increasing engraftment of *P. copri* type strain (n = 12 per group). **a** TCST. **b** LDB test. **c** YM test. **d** EPM test. **e** OFT. **f** MWM test. Data are represented as mean ± SEM. ** *p* < 0.01, ns depicts no significant difference.

**Fig. S6.**
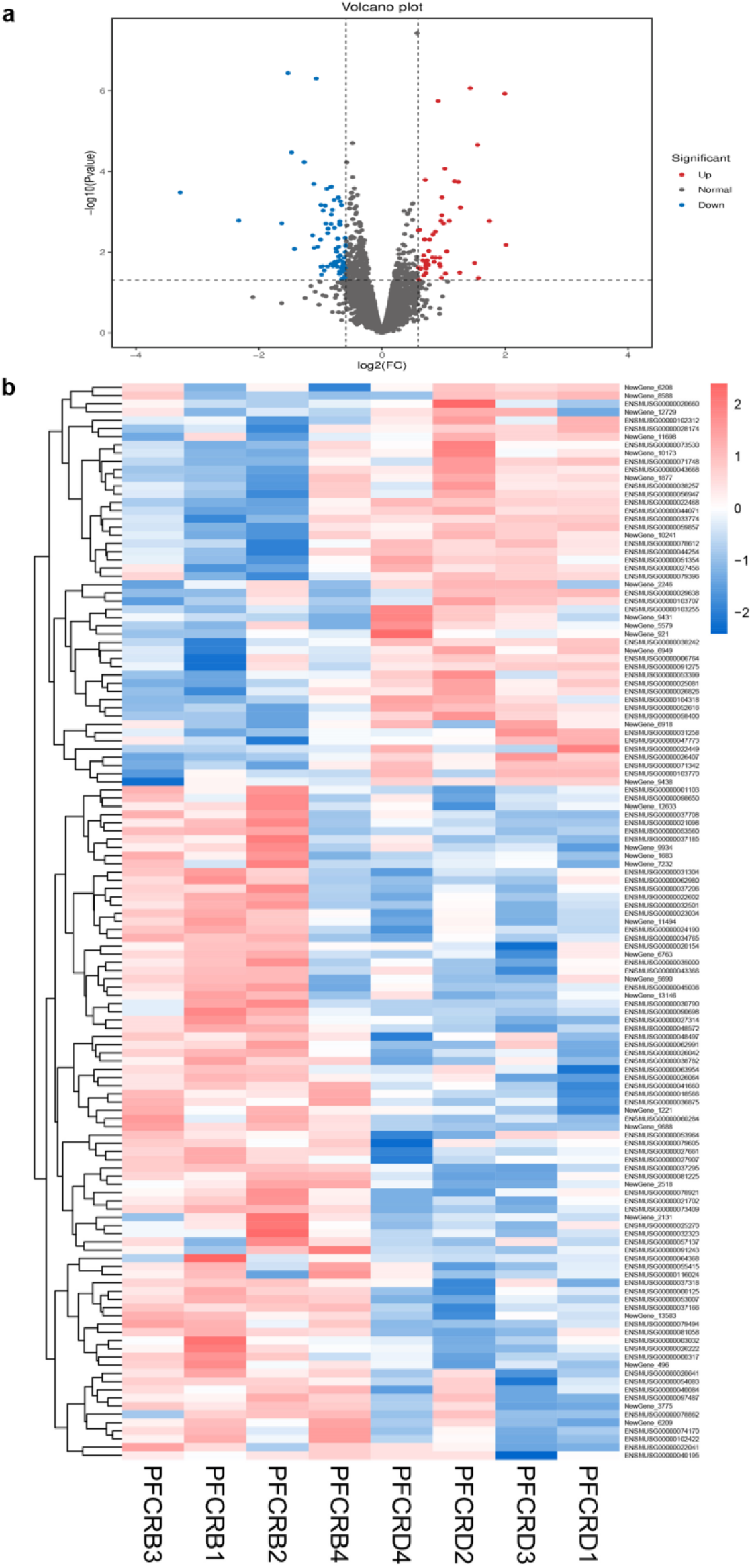
Differentially expressed genes in PFC after increasing engraftment of *P. copri* type strain (n = 4 per group). **a** Volcano plot with thresholds of fold change (FC) ≥ 1.5 and false discovery rate (FDR) adjusted p-value (q-value) < 0.05. **b** Scaled relative expression (fragments per kilobase of transcript per million fragments mapped, FPKM) of differential genes.

**Fig. S7.**
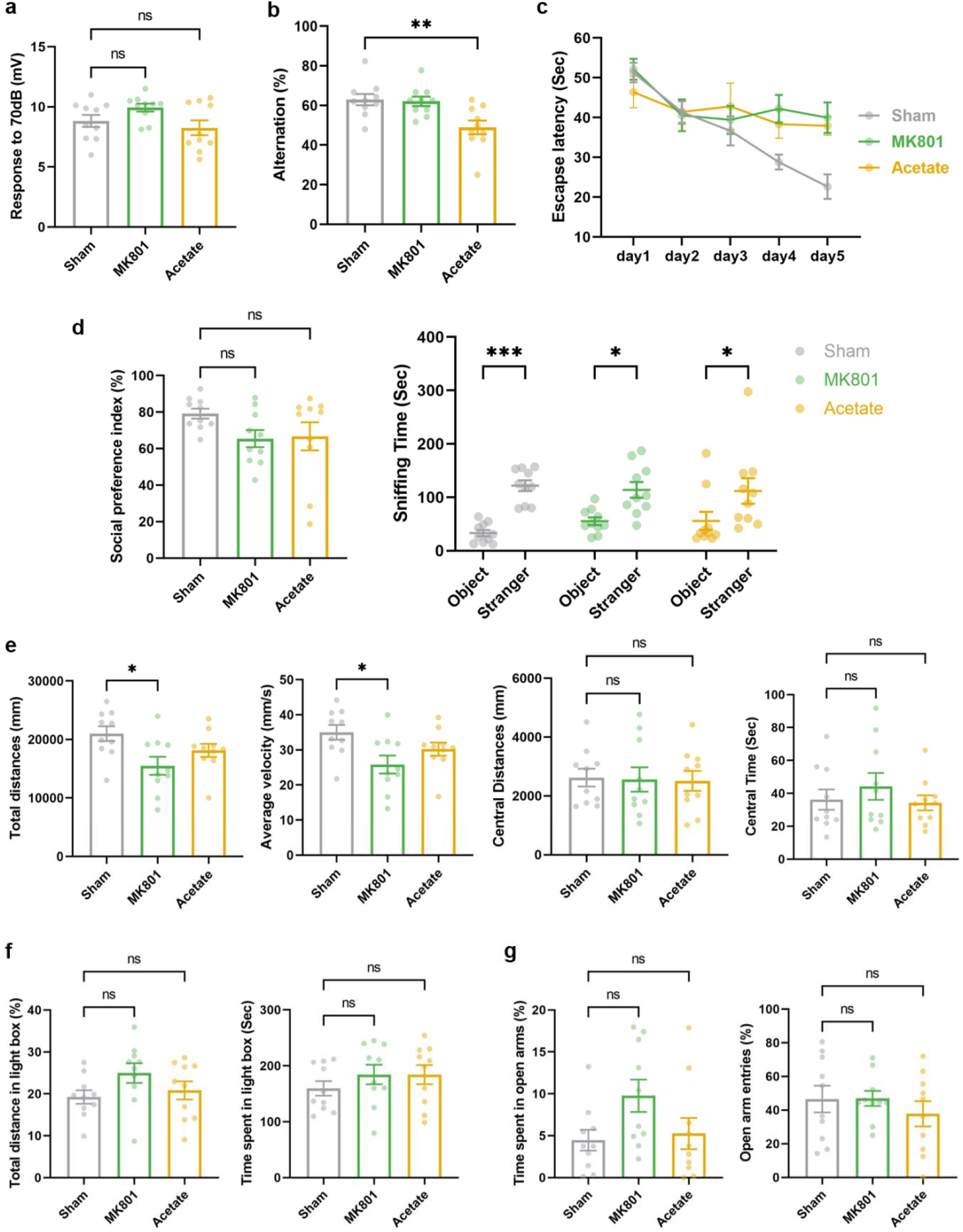
Behavioral experiments after increasing acetate intake (n = 10 per group). a PPI test. b YM test. c MWM test. d TCST. e OFT. f LDB test. g EPM test. Data are represented as mean ± SEM. * *p* < 0.05, ** *p* < 0.01, *** *p* < 0.001, ns depicts no significant difference.

